# Personalized Pediatrician-Scientist Training Program Optimizes Institutional Research Investment

**DOI:** 10.64898/2026.09.21.26363577

**Authors:** Erin J. Plosa, Mark Denison, Kemberlee Bonnet, Anika Yarlagadda, Nikita Muthakana, David G. Schlundt, Julie A. Bastarache, Michael R. DeBaun

## Abstract

**Objective:** Physician-scientists are a diminishing subset of the pediatrician work force, uniquely trained to advance child health through scientific discovery. The current financial landscape of academic medical centers makes traditional resource-intensive training programs untenable.Continued renewal of this critical pediatric workforce requires new resource-efficient methodologies. Thus, we evaluated the impact of the NICHD supported Vanderbilt K12 program, a personalized, intensive training program.

**Study Design:** We tracked academic outcomes for scholars with K12 appointments between January 2015 and December 2022. Our primary outcome was receipt of a mentored career development award (K08 or K23). Successful transition from mentored to independent NIH funding (K to R transition) was used as a secondary outcome. Academic outcomes for faculty participating in a large, institutional training program, the Vanderbilt Faculty Research Scholars program (VFRS), served as a comparison cohort. A complementary qualitative evaluation of the K12 program was performed.

**Results:** K12 scholars earned career development awards (73%) at an equivalent rate to VFRS faculty (71%) and transitioned K to R funding at equivalent rates, 45% of K12 scholars compared to 50% for VFRS. Qualitative analysis demonstrated K12 scholars valued a personalized training program that incorporated self-efficacy and adaptability, key tenets of Social Cognitive Career Theory.

**Conclusion:** Personalized physician-scientist training programs may enhance efficient utilization of research resources in Pediatric academic departments and serve as a viable alternative to traditional programs. Our qualitative evaluation suggests scholar self-efficacy and adaptability can be achieved through a personalized training program.

## Introduction

As a workforce, physician-scientists are uniquely positioned to apply scientific discoveries to clinical questions, thereby facilitating advancements in human health. Historically, physician-scientist innovation accounts for 20% of medical device patents in the United States (1), despite comprising only 1.3% of the physician workforce (2). Over the past two decades, the total number of physician-scientists in the U.S. has steadily declined, while the average age has increased, resulting in a shrinking physician-scientist workforce without adequate renewal (2).

The physician-scientist training pathway is lengthy, requiring both personal commitment and significant institutional investment. Physician-scientist career development has traditionally centered around large institutionally supported training programs, consisting of didactic seminars, one-on-one scientific mentorship, and institutional research support. The current financial landscape of academic medical centers threatens the viability of this career pathway. Thus, new programmatic methodologies that efficiently utilize academic resources are needed to support early career physician-scientists. To address this gap, we developed a new training program structure based on key tenets of the Social Cognitive Career Theory (3). This conceptual framework explains how individuals choose career paths, maintain motivation, and achieve career goals through perceptions of self-efficacy. Applicable to early career physician-scientists, our training program sought to enhance scholar self-efficacy and adaptability through personalized mentorship.

## Methods

According to the Vanderbilt University Medical Center Institutional Review Board (IRB) guidelines, this project did not constitute human subjects research by meeting quality improvement criteria; therefore, formal IRB review was not required. Supported by the Vanderbilt Department of Pediatrics, our personalized physician-scientist training program provided intensive support for three concurrent early career physician-scientists as they prepared competitive mentored career development awards. The Vanderbilt Department of Pediatrics and the NICHD K12 award, with a total annual budget of $410,522, provided salary support for program leadership, program participants (80% protected time for participants), and a modest research stipend of $25,000 annually to each scholar. K12 participants were selected by a competitive application process, thereafter, referred to as K12 scholars. All mentors of the K12 scholars were independent investigators with current NIH funding. In this quality improvement project, we tested the primary hypothesis that ***participation in a departmentally supported intensive, personalized training program would result in mentored and independent research award funding success rates equivalent to participation in a traditional, institutionally supported training program (the Vanderbilt Faculty Research Scholars program, or VFRS)***.

We defined academic success by primary and secondary outcomes: 1) receipt of a mentored career development award (K08, K23) and 2) receipt of an independent award (R01 or equivalent). Publicly available data on funding and publications was obtained through NIH RePORTER and PubMed. For two group comparisons, Fisher’s exact test compared categorical data and Student’s t-test compared quantitative data, utilizing Prism GraphPad v8 (Boston, MA).

### The K12 personalized training program focused on self-efficacy and adaptability in pediatric physician-scientists

A major goal of our personalized K12 training program was to improve scholar self-efficacy, the belief in one’s own ability to achieve valued goals. We chose this focus based on the University of California San Diego’s National Center of Leadership in Academic Medicine (NCLAM) report that demonstrated participation in a formal mentorship program improved junior faculty self-efficacy (4), and that self-efficacy directly related to increased academic productivity (4, 5). We targeted scholar self-efficacy through formation of a personalized monthly mentoring group known as the “K12 Club.” Topics for the monthly K12 Club meetings were selected to address scholar concerns as they arose, limiting unnecessary delays in research progress. The training directors contacted each scholar in the week prior to K12 Club to assess their current needs and progress, subsequently selecting topic(s) that assisted them in formulating ***scholar-driven solutions*** (**Figure 1**). Example scholar challenges include uncertainties about how to build a team, managing sunk costs, and feelings of isolation or marginalization. The personalized K12 Club curriculum included group discussion, book review, and personal goal accountability, as well as served as a platform for near-peer mentorship. Active listening by the training directors, application of a flexible training curriculum, and the provision of actionable tools directly enhanced scholar self-efficacy, while implementation of scholar-driven solutions improved adaptability.

**Figure 1.**
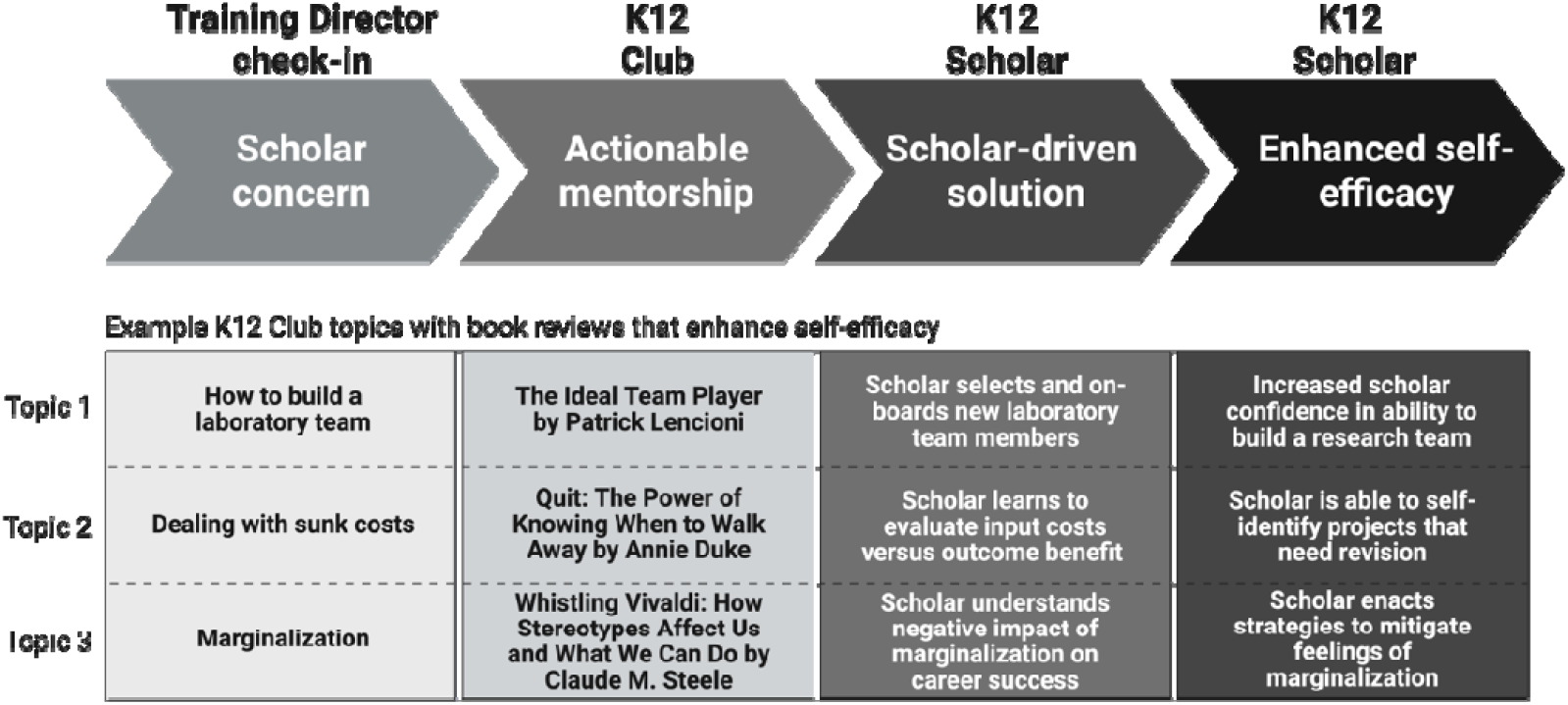
Personalized Pediatric Physician-Scientist Training Program Enhances Scholar Self-Efficacy. Created in BioRender. Plosa, E. (2026) https://BioRender.com/0ai25n2

### Comparison of K12 to VFRS outcomes evaluate departmentally versus institutionally supported physician-scientist training programs

We evaluated the quality of the K12 program by comparing the academic outcomes of the K12 scholars to faculty participating in the VFRS, an institutionally supported training program with an established “gold standard” track record of success. The VFRS training program is administered through the Vanderbilt Institute for Clinical and Translational Research (from a total annual budget of $10,726,281), which supports both physician-scientists (MD and MD, PhD degrees) and clinician-scientists (faculty with PhD degrees working in a clinical department). We included all K12 and VFRS faculty appointed to their respective programs between January 1, 2015 and December 31, 2022 in the analysis. We tracked scholarly outcomes for a minimum of three years from training program appointment (through December 31, 2025). The K12 supported three scholars concurrently for up to 36 months, resulting in 11 faculty with three years of follow up available for outcome analysis. Similarly, the VFRS provided up to 36 months of academic support, with 17 concurrent positions, resulting in 53 faculty with three years of follow up for our analysis, 28 of whom were physician-scientists and 25 clinician-scientists. The K12 and VFRS scholars received equivalent salary and research support. The VFRS program included a monthly career development seminar series and personalized, intensive training but the training curriculum did not address scholar resilience and adaptability.

### Qualitative analysis was performed as a complementary method of iterative K12 program improvement

In addition to quantitative evaluation, we sought to evaluate the success of the K12 program through qualitative analysis to gain an in-depth understanding of the program strengths and weakness, allowing for contextual relevance and a holistic scholar perspective. In collaboration with the Vanderbilt University Qualitative Research Core (VU-QRC), we created a semi-structured interview guide (**Supplemental Figure 1**). Interviews were independently conducted over the telephone by study personnel trained in qualitative methods (K.B., MA in Social Psychology, VU-QRC Senior Research Manager, female, 14 years qualitative research experience, no prior relationship with study participants). Interviews were audio recorded, transcribed verbatim using an IRB-approved transcription service (rev.com), and de-identified to preserve confidentiality. Interview responses were qualitatively coded by two co-authors (KB and DS) using a hierarchical coding system. To ensure consistency in applying the codes, intercoder reliability was achieved through reconciliation discussion sessions among coders. The coded data were analyzed in accordance with established criteria for reporting qualitative research guidelines (6). A conceptual framework was created by an iterative deductive/inductive analysis of the deidentified coded responses (7-9). Deductively, we were guided by Social Cognitive Career Theory, which describes individuals as intentional agents driven by the need to achieve their goals by exercising autonomy and having a sense of control over their lives.

Inductively, our analysis consisted of using the coded quotes to identify higher-order themes and relationships among themes. We contacted twelve current or former K12 scholars (one current scholar was included in the qualitative analysis that did not meet follow up criteria for quantitative analysis). Eleven of twelve K12 scholars participated in the semi-structured interviews, providing qualitative data in seven major coding categories: program strengths, program challenges, program improvement, grant preparation, mentorship training, program recommendation to others, and career trajectory. We analyzed the coded response frequency and their hierarchical categorization to identify the most important themes related to strengths and challenges of the K12 program.

## Results

### VFRS comparison demonstrates equivocal success rates for the K12 program

Our quantitative analysis compared the academic outcomes of K12 and physician-scientist VFRS scholars (ps-VFRS) (**Table 1**). For our primary outcome, the K12 and ps-VFRS scholars exhibited equivalent rates of mentored career development awards (73% of K12 scholars and 71% for VFRS, *p* = 0.69), with both cohorts of scholars earning mentored awards in equivalent amounts of time (K12 scholars in 2.2 years (SD 0.7 years) compared to 1.6 years (SD 1.5 years) for ps-VFRS scholars, *p* = 0.24). Our secondary analysis also demonstrated equivalent success rates in obtaining independent research funding with 45% of K12 scholars and 50% of ps-VFRS scholars obtaining independent funding, *p* > 0.99. Similarly, the K12 and ps-VFRS scholars published similar numbers of manuscripts per year. Our analysis considered the physician-scientists VFRS subgroup only (MD and MD, PhD), as directly comparable to the physician-scientist group of K12 scholars. Data for the VFRS clinician-scientist PhD faculty is reported here as well (**Supplemental Figure 2**). Taken together, these findings indicate that a departmentally sponsored personalized training program equivalently supports physician-scientist career development as compared to a large, institutionally sponsored program.

**Table 1.**
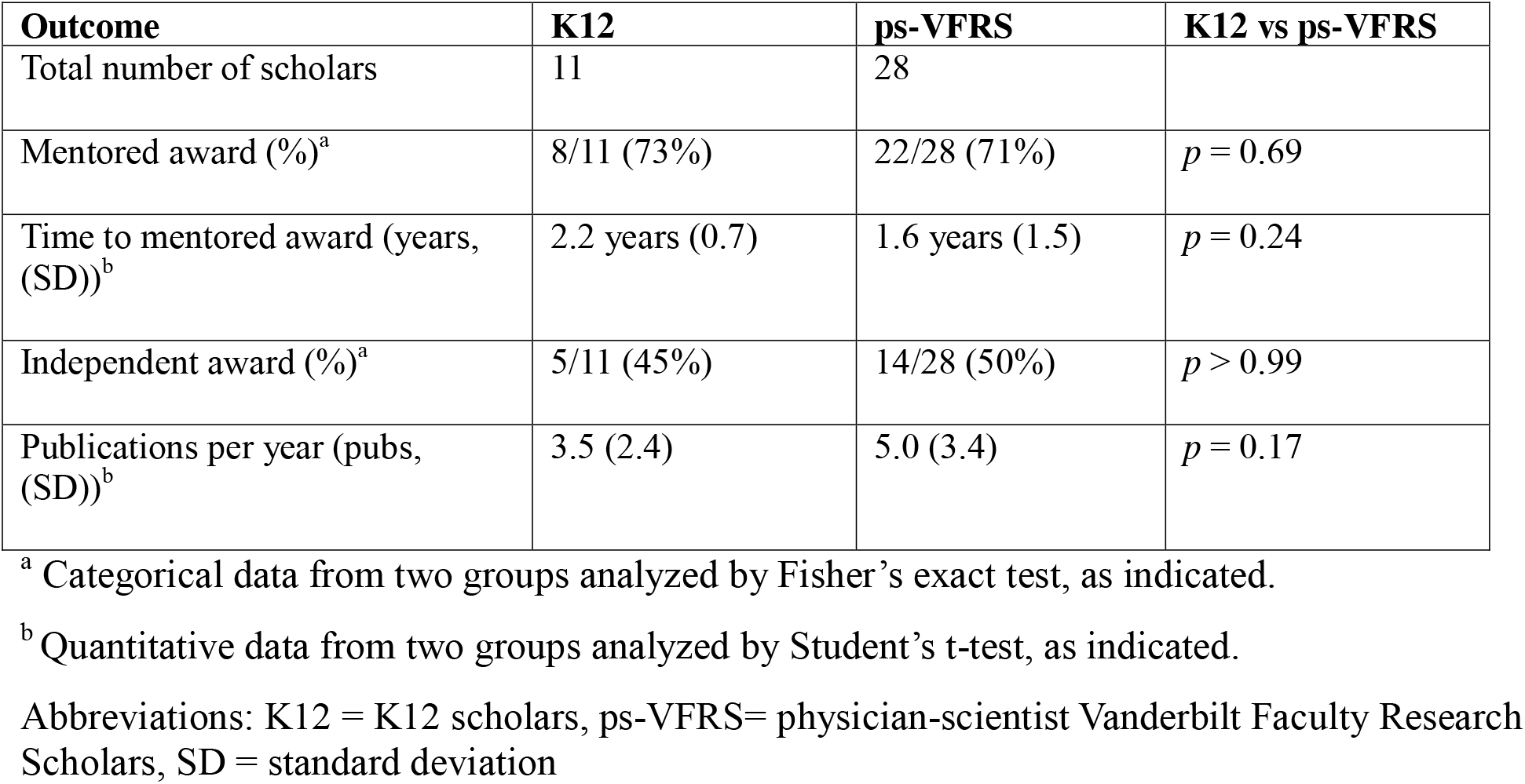
Analysis of Academic Outcomes for K12 and Physician-Scientist VFRS Scholars.

| **Outcome** | **K12** | **ps-VFRS** | **K12 vs ps-VFRS** |
| --- | --- | --- | --- |
| Total number of scholars | 11 | 28 |  |
| Mentored award (%)<sup>a</sup> | 8/11 (73%) | 22/28 (71%) | $p = 0.69$ |
| Time to mentored award (years, (SD))<sup>b</sup> | 2.2 years (0.7) | 1.6 years (1.5) | $p = 0.24$ |
| Independent award (%)<sup>a</sup> | 5/11 (45%) | 14/28 (50%) | $p > 0.99$ |
| Publications per year (pubs, (SD))<sup>b</sup> | 3.5 (2.4) | 5.0 (3.4) | $p = 0.17$ |

<sup>a</sup> Categorical data from two groups analyzed by Fisher's exact test, as indicated.
<sup>b</sup> Quantitative data from two groups analyzed by Student's t-test, as indicated.
Abbreviations: K12 = K12 scholars, ps-VFRS= physician-scientist Vanderbilt Faculty Research Scholars, SD = standard deviation

**Figure 2.**
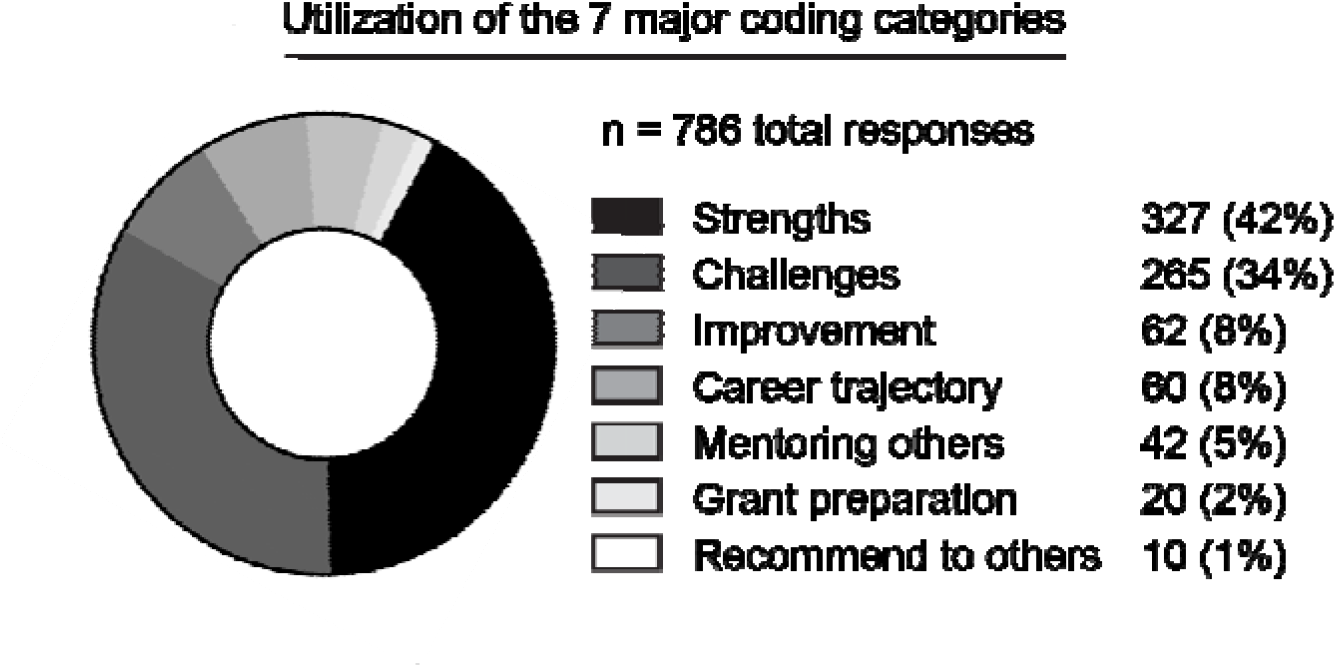
Qualitative thematic analysis demonstrates utilization of seven major coding categories with strengths outnumbering challenges.

### Qualitative analysis demonstrates early career physician-scientists value a personalized training program

Our qualitative analysis contained 786 separate coded responses from eleven K12 scholars. Of the seven major response domains, 327 (42%) categorized as program strengths, the category with the highest frequency (**Figure 2**). Hierarchical analysis clustered 14 program strengths into just three subcategories, indicating widely shared perceptions of program strengths among the K12 scholars (**Figure 3**). Notably, all eleven K12 scholars (100%) discussed the value of personalized mentoring by the training directors, with eight of eleven (73%) scholars providing detailed accounts of personal growth, increased resilience, and improved adaptability (**Supplemental Figure 3**).

**Figure 3.**
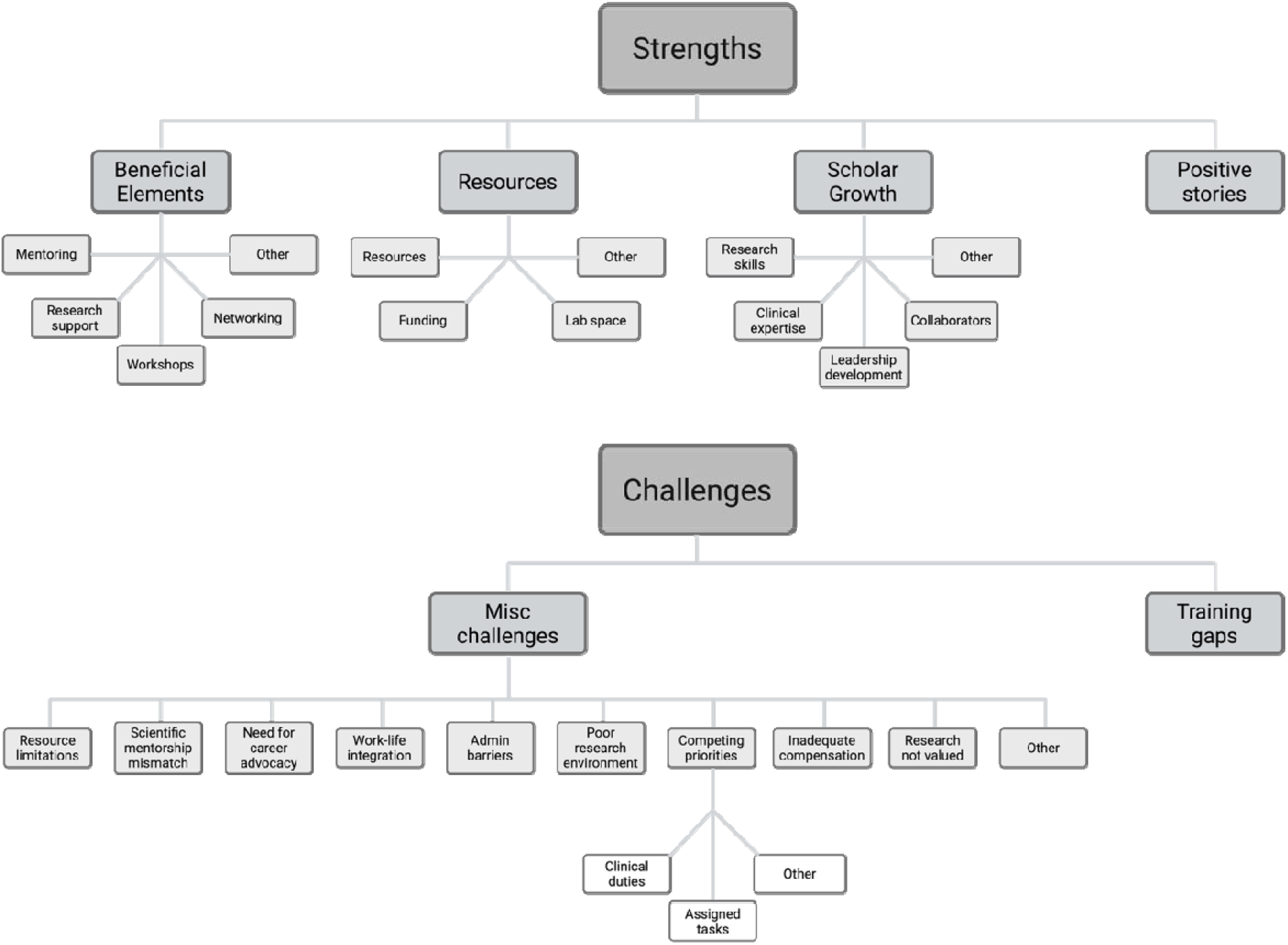
Hierarchical analysis of coded responses demonstrates widely shared perceptions of K12 program strengths and individual challenges.

Networking was the second most frequently cited strength of the K12 program. Scholars described local networking within the K12 Club (near-peer mentorship) that normalized setbacks and improved resilience, institutional networking through training directors that improved accessibility to research resources, and national networking through training directors that resulted in new collaborations (Supplemental Figure 3).

The K12 scholars discussed a series of individual challenges that did not cluster thematically (Figure 3, Supplemental Figure 3). Resource limitations and competing priorities were the most frequently discussed challenges. The individual nature of these challenges suggests a personalized approach to physician-scientist training programs would benefit scholars.

In summary, independent thematic analysis identified program improvement over time and that multiple facets of the K12 program structure accelerated research progress through parallel approaches. These findings support the concept that a personalized physician-scientist training program with iterative program evaluations enhances scholar academic success, as well as efficient utilization of existing departmental and institutional resources.

## Discussion

To our knowledge, our report is the first evaluation of a physician-scientist training program based on Social Cognitive Career Theory with comparison to a large, traditional institutional training program. Our mixed-methods approach suggests that intentional personalized programming targeting scholar self-efficacy and adaptability facilitates academic success as defined by receipt of a mentored career development award. Our findings build on prior studies that reported scholar research self-efficacy is positively associated with increased intention to pursue or remain in a research career (10, 11), as well as positively associated with increased number of grant submissions (12, 13). Since self-efficacy and adaptability mitigate feelings of marginalization, training methodologies that improve these traits may prove particularly valuable in the future. A recent study by Shalev et al of NIH K-awardees across multiple institutes suggests that early career researchers perceive declining career stability, as well as decreased likelihood of remaining in a research career or submitting an R01 application (14). Consistent with the concept of self-efficacy as an antidote for career marginalization, our qualitative results indicate that eight of eleven K12 Scholars described personal growth, increased resilience, and improved adaptability having participated in a program intentionally focused on scholar self-efficacy and adaptability. Taken together, our findings support the inclusion of Social Cognitive Career Theory concepts into physician-scientist training programs as a potentially modifiable approach to strengthen research career intention and promote physician-scientist workforce retention.

Personalization of the K12 Club curriculum is a distinctive feature of the Vanderbilt K12 program. Rather than following a pre-determined set curriculum, K12 Club monthly topics addressed scholars concerns in-the-moment. This framework facilitated timely mentoring by the Training Directors and, importantly, development of scholar-driven solutions with immediate applicability. The variability in scholar challenges identified by our qualitative study supports a personalized approach for a physician-scientist training curriculum. Lack of thematic clustering of scholar challenges suggests a set curriculum would less efficiently address scholar concerns.

Alignment of barriers to solutions in recurring monthly discussions repeatedly demonstrates the value of adaptability to the scholars, thereby further enhancing scholar self-efficacy.

Our findings suggest that a departmentally based personalized model may be a viable alternative to large institutionally supported physician-scientist training program. As a primary outcome, we observed similar rates in the receipt of mentored career development awards for K12 scholars compared with VFRS participants, despite distinctly different program structures. We demonstrated successful physician-scientist development may not require reproducing every component of a large, centralized training program. Rather, resource-efficient elements of the Vanderbilt K12 Program could be directly implemented by smaller or emerging physician-scientist training programs. Personalized monthly meetings, near-peer mentoring, and an emphasis on timely scholar-driven solutions are resource-efficient and transferable features. The applicability of the K12 Club program has broader implications for addressing the pediatrician- scientist workforce deficit. Our study describes a successful physician-scientist training program model that could be applied to a range of small to large programs, without dependence on substantial institutional programs in place.

Our K12 program quality improvement project has several limitations related to small numbers of participants. The K12 quantitative cohort contained only 11 scholars versus 28 ps- VFRS scholars and assignment was determined by application and appointment to each program, rather than randomization. The independent qualitative analysis was a necessary step given the personalized nature of our program but was completed only for the K12 cohort. However, since a qualitative analysis of ps-VFRS scholars is not available, our results support the feasibility and promise of the personalized model but cannot establish that personalization caused the outcomes or that the programs are truly equivalent.

In summary, our evaluation suggests that a personalized approach to physician-scientist career development promotes efficient utilization of research resources with equivalent academic outcomes to larger, institutionally supported programs. Our study demonstrates value in incorporating key tenets of Social Cognitive Career Theory to physician-scientist training programs. These findings are applicable to academic institutions as an approach to maximize utilization of research investments and may help address the pediatrician-scientist workforce deficit.

## Data Availability

All data produced in the present study are available upon reasonable request to the authors.

## Abbreviations

(IRB): Institutional Review Board
(NICHD): Eunice Kennedy Shriver National Institute of Child Health and Human Development
(VRFS): Vanderbilt Faculty Research Scholars Program
(NCLAM): National Center of Leadership in Academic Medicine.

## Ethical approval

Not applicable due to being a Quality Improvement study design.

## Data statement

The data that support the findings of this study are available on request from MRD.

## Acknowledgements

The authors would like to thank the Vanderbilt K12 Scholars, the Vanderbilt Faculty Research Scholars Program, and the Vanderbilt Qualitative Core.

## AI statement

During the preparation of this work, the author(s) used ChatGPT and Claude in a supportive capacity for proofreading of this manuscript. AI technology was not used in a generative capacity. After using these tools, the authors reviewed and edited the content as needed and take full responsibility for the content of the published article.

