## Supplemental Figures for "Personalized Pediatrician-Scientist Training Program Optimizes Institutional Research Investment"

**Supplemental Figure 1:** Semi-structured Interview Guide for Pediatric NICHD K12 Scholars in the Vanderbilt Department of Pediatrics

**Objective:**
To gather valuable insights into the strengths and areas for improvement of the Pediatric NICHD K12 program, fostering continuous growth and improved scholar experience.

**Section 1: General Information**
1. Name (optional):
2. Date of Interview:
3. Start Date of Program:
4. Completion Date of Program:
5. Primary Mentor(s):
6. Research Focus Area:

**Section 2: Program Strengths**

1. What aspects of the Pediatric NICHD K12 program did you find most beneficial for your career development?
- Mentorship
- Research support
- Didactic sessions/workshops
- Networking opportunities
- Grant writing and funding guidance

2. Which program resources were most valuable to you?
- Lab space
- Funding
- Administrative support

3. How did the program contribute to your growth as a clinician-researcher?
- Research skills
- Clinical expertise
- Leadership development
- New connections leading to collaborations

4. Can you describe any specific moments or achievements that you felt the program helped you achieve? What particular features of the program that facilitated your success?

**Section 3: Program Challenges and Areas for Improvement**

1. What challenges did you face during your time in the program?
- Limited research resources
- Inadequate scientific mentorship
- Inadequate career mentorship
- Insufficient career sponsorship/ advocacy by mentors
- Insufficient career sponsorship/ advocacy by leadership
- Insufficient support for work-life balance
- Administrative hurdles
- Non-collaborative research environment
- Research time limited by clinical duties by quantity (call, service, clinic)
- Research time limited by clinical duties by timing/ schedule (call, service, clinic)

- Research time limited by additional assigned tasks (teaching, interviews, clinical guidelines or committees)
- Imbalance between workload and financial compensation
- Research program not valued by colleagues
- Research program not valued by mentors
- Research program not valued by leadership

2. Were there gaps in the curriculum or training that you believe should be addressed for future scholars?

3. What improvements could be made to the program mentorship structure?
- Frequency or timing of mentor meetings
- In-person vs virtual mentor meetings
- Quality of feedback
- Accessibility

4. Did you feel adequately prepared for grant submissions and career advancement? If not, what areas need more focus?
- Timeline preparation
- Scientific writing
- Application instructions/ mechanics of submission
- Examples of successful applications

5. Were there any logistical or administrative aspects of the program that you found particularly challenging?
- Adherence to 80% protected time
- Timing/ amount of clinical work limited participation in program

**Section 4: Program Impact and Career Outlook**

1. How has the program influenced your career trajectory?
- Academic position
- Research independence
- Funding opportunities

2. What are your career goals post-program, and how well did the program prepare you to achieve them?

3. Would you recommend the Pediatric NICHD K12 program to colleagues? Why or why not? Were there any unanticipated negatives to participating in the program?

**Section 5: Additional Feedback**

1. What additional resources or opportunities would have enhanced your experience?

2. Do you have any suggestions for improving the overall scholar experience?

3. Is there anything else you'd like to share about your time in the program?

4. How many trainees have you mentored (defined as primary research mentor for > 6 weeks). What awards have they won?
    - Trainees- summer students (> 6 weeks), undergraduate students, gap year students, medical students, graduate students, residents, clinical fellows, post-docs
    - Abstracts selected for oral presentation at national/ international meetings, travel awards, poster awards, young investigator awards, foundation grants, K08, K23, R03, R01, or similar

**Thank you for your time and valuable feedback! Your input will help shape the future of the Pediatric NICHD K12 program at Vanderbilt.**

**Supplemental Figure 2.** Academic Outcomes for K12 and VFRS Scholars, by physician-scientist and clinician-scientist subgroup.

| **Outcome** | **K12** | **VFRS** | **K12 vs VFRS** |
|---|---|---|---|
| **Total number of scholars** | 11 | 53 | |
| MD (%) | 5/11 (45%) | 17/53 (32%) | |
| MD, PhD (%) | 6/11 (55%) | 11/53 (21%) | |
| PhD (%) | | 25/53 (47%) | |
| **Mentored Award** (%)[a] | | | |
| Physician-scientist | 8/11 (73%) | 22/28 (71%) | $p = 0.69$ |
| Clinician-scientist | | 11/25 (44%) | |
| **Time to mentored award** (years, (SD))[b] | | | |
| Physician-scientist | 2.2 years (0.7) | 1.6 years (1.5) | $p = 0.24$ |
| Clinician-scientist | | 0.4 years (1.0) | |
| **Independent award** (%)[a] | | | |
| Physician-scientist | 5/11 (45%) | 14/28 (50%) | $p > 0.99$ |
| Clinician-scientist | | 12/25 (48%) | |
| **Publications per year** (SD)[b] | | | |
| Physician-scientist | 3.5 (2.4) | 5.0 (3.4) | $p = 0.17$ |
| Clinician-scientist | | 4.3 (2.9) | |

[a] Categorical data from two groups analyzed by Fisher's exact test, as indicated.

[b] Quantitative data from two groups analyzed by Student's t-test, as indicated.

Definitions: Physician-scientist = scholars with MD or MD, PhD degrees, Clinician-scientists = PhD faculty appointed to a clinical department, K12 = K12 scholars, VFRS = Vanderbilt Faculty Research Scholars, SD = standard deviation

**Supplemental Figure 3.** Qualitative Analysis demonstrates personalized mentoring is valued by K12 scholars.

> *I think the two things that stand out the most are, one, absolutely mentorship from Dr. DeBaun and Dr. Denison. The two of them,* ***their expertise, their candor is by far one of the best things about that program to me****. They both definitely take the time, they're very* ***thoughtful****, they're* ***honest****, they're open, they're open to talking about literally anything that we want to talk about and* ***push us to think about things that we may not realize we need to think about****. (Participant 1)*

> *I know that we read a couple of* ***articles on women in medicine*** *and women in science, and those were good. Those were good. Those discussions were always quite spirited and engaging, and* ***I think really led to a lot of growth from my standpoint****, just in terms of being* ***exposed to so many different opinions*** *on things that are so important in day-to-day, both clinical and research leadership. I found that that was helpful. (Participant 2)*

> *We would discuss chapters, how to be* ***resilient****, how to* ***manage time****, how to* ***set up your lab****, how to* ***manage your lab****, man management, time management. ... But definitely* ***how to stay positive****, how to manage time,* ***how to write grants****, how to set up a lab, how to take the lab to the next level. Those are some of the lessons discussed and learned from each other as well. (Participant 3)*

> *I had lots of moments where I wasn't successful and that's fine. I think when I was younger, I viewed that as something really bad and scary. But I think along the road all*

*these mentors have shown and told me and been like, "you know how many times we've failed at things or not gotten some the things?" That's just what happens. But you know what? If you keep trying and you keep doing those things, you could do what you want to do. (Participant 5)*

Qualitative Analysis: Networking is a Strength of the K12 Training Program

*Dr. DeBaun and Dr. Denison has helped me make connections and even some potential collaborations which will be, especially in the current funding situation, essential for me being able to be successful in the future. (Participant 1)*

*Again, through this* ***peer-to-peer*** *thing* [K12 Club]*, I was* ***able to identify a resource that existed within in my building****, in my hall that I didn't even know was there until we chatted. And I was able to generate a huge piece of data just because I had talked to another one of the K12 people. (Participant 2)*

[Regarding the K12 Club] *Having a soundboard and other people who are working really hard and even struggling at times around the same things is refreshing to be like, okay, we're in this together. And then the layers of mentors that have then been like, "we have the exact same issues" or "I also struggled with that" makes it attainable and gives me still hope being like they also had these issues and challenges and they're doing quite well and they've worked hard. So, it gives me still a little bit of energy to keep going and trying. (Participant 5)*

Qualitative Analysis: Individual Scholar Challenges

*I also was in a situation where we were frequently understaffed within our division. And so often times, if there was help needed, I was kind of the person to ask. And it was one of those where there's a constant pull between the responsibilities as a scientist and your desire to support your division and colleagues. And so I would pick up shifts, but that, again, meant I wouldn't be in the lab. (Participant 2)*

*So I mean, there's clearly the recipe for success that exists in the department, but in some ways, one size doesn't fit all. There are some concepts about establishing your lab that are kind of universal, and I think that the K12 group does really well with those, but I think that there's an opportunity for improvement in flexibility and acknowledging differences in different areas of science and as that relates to ability and/ or willingness to be flexible in the mentorship. (Participant 9)*
